# Non-invasive Diagnosis of Deep Vein Thrombosis from Ultrasound with Machine Learning

**DOI:** 10.1101/2021.01.23.21249964

**Authors:** Bernhard Kainz, Antonios Makropoulos, Jonas Oppenheimer, Christopher Deane, Sven Mischkewitz, Fouad Al-Noor, Andrew C Rawdin, Matthew D Stevenson, Ramin Mandegaran, Mattias P. Heinrich, Nicola Curry

**Author notes:** **Corresponding Author: Bernhard Kainz, PhD**. **Co-Authors:**.

## Abstract

Deep Vein Thrombosis (DVT) is a blood clot most found in the leg, which can lead to fatal pulmonary embolism (PE). Compression ultrasound of the legs is the diagnostic gold standard, leading to a definitive diagnosis. However, many patients with possible symptoms are not found to have a DVT, resulting in long referral waiting times for patients and a large clinical burden for specialists. Thus, diagnosis at the point of care by non-specialists is desired.

We collect images in a pre-clinical study and investigate a deep learning approach for the automatic interpretation of compression ultrasound images. Our method provides guidance for free-hand ultrasound and aids non-specialists in detecting DVT.

We train a deep learning algorithm on ultrasound videos from 246 healthy volunteers and evaluate on a sample size of 51 prospectively enrolled patients from an NHS DVT diagnostic clinic. 32 DVT-positive patients and 19 DVT-negative patients were included. Algorithmic DVT diagnosis results in a sensitivity of 93.8% and a specificity of 84.2%, a positive predictive value of 90.9%, and a negative predictive value of 88.9% compared to the clinical gold standard.

To assess the potential benefits of this technology in healthcare we evaluate the entire clinical DVT decision algorithm and provide cost analysis when integrating our approach into a diagnostic pathway for DVT. Our approach is estimated to be cost effective at up to $150 per software examination, assuming a willingness to pay $26 000/QALY.

## Introduction

Venous thromboembolism (VTE) is associated with a major global burden of disease. Worldwide, the incidence of VTE is 1 – 3 per 1000 individuals, rising to 2 – 7 per 1000 in individuals aged over 70 years, and 3 – 12 per 1000 in those over 80 years^1^. VTE, DVT and pulmonary embolus are the leading cause of hospital related Disability-Adjusted Life Years lost^2^.

Using these estimates, and using the most conservative incidence figure, globally at least 7.7 million people will require investigation for VTE every year. An aging population across many countries will lead to a greater health burden, particularly in middle- and low-income countries where early death from is decreasing. Mortality from VTE is common, a European study estimated 534 000 deaths per year^3^ and a similar study in the US reported 300 000 deaths per year^4^. Deep vein thrombosis (DVT) has a high level of morbidity and 30 - 50% of the surviving patients develop long-term symptoms in their affected leg (post-thrombotic syndrome)^5^.

In high income countries, the routine practice to diagnose patients after a positive D-dimer blood test and an indicative evaluation using the Wells score^6^ is to confirm or rule out a suspected DVT with a two- or three-point ultrasound scan. Ultrasound scans are most commonly performed in a radiology department of a hospital by a highly trained radiographer/radiologist.

Currently, no reliable test is available that can be used in a general healthcare setting (GP practice, community hospital, on a hospital ward) or be used remotely at the point of care (nursing home, patient’s home). Between 85 – 90% of patients presenting to their GP in high income countries with a suspected DVT will be investigated only to find no evidence of a thrombus^5^. Many patients will receive unnecessary anticoagulants with many potential side-effects through an often painful subcutaneous injection whilst waiting more than the recommended four hours for their scan. Safely negating this wait would improve patient satisfaction, reduce the burden of high-risk treatment (anticoagulants confer haemorrhagic complication risks) and reducing healthcare costs. Rapid diagnosis is known to improve compliance to regulatory guidelines that state DVT should be diagnosed within 24 hours^6-8^.

Clinical evidence that DVT examinations can be performed by nurses has been shown by^9-11^. However, confidence in acquiring ultrasound images is generally low because of the required image interpretation skills and liability concerns, which inhibits wide-scale adoption of such approaches.

In this study we evaluate if Machine Learning (ML) technology can provide anatomical image acquisition guidance and point of care diagnostic support. Such ML technology is currently often summarised as Artificial Intelligence (AI) support systems.

Our hypothesis is that ML technology can complement the clinical pathway and provide front line of care personnel with the necessary confidence and skills to perform ultrasound DVT screening autonomously. Early modelling has been undertaken to assess the potential cost-effectiveness of such an approach.

## Materials and Methods

### Study design

This study is a primary analysis of compression ultrasound screen recordings performed on prospectively enrolled patients at the Oxford Haemophilia and Thrombosis Centre adult DVT clinic. The University of Oxford, UK, approved the study (Ethics: 18/SC/0220, IRAS 234007). All participants provided written informed consent. Eligible participants were consecutively recruited between January 2019 and December 2019. Patients were approached about participation in the study after their routine ultrasound DVT examination. After study information and consent, they were scanned for a second time by an expert radiographer. During the second scan a mobile ultrasound device was used (Clarius L7 (2017) or Philips Lumify L7). Built-in functionality was used to record the examination as mp4 videos. Patient identifying information has not been recorded in the videos but separately in a spreadsheet where it was tagged with a unique identifier (UID) by co-author Ch.D.. Only the UID was used during downstream analysis.

In this work we call the data set from the Oxford Haemophilia and Thrombosis Centre the *external validation set*.

Since the analysed prototype device is based on a ML computer algorithm, training data and preliminary testing data is required. Thus, preliminary data acquisition was performed on healthy volunteers (n=246) and nine consenting DVT patients. Acquisition has been performed by two radiologists and three trained engineers. We call this algorithm training data *training set* (Table A1 in Appendix A). The volunteers and patients that have been left out from training to monitor the algorithm’s performance during development are included in the *internal validation set* (Table A2 in Appendix A).

Image quality control has been performed by a medical student according to a specialist-defined scheme (Appendix C).

All compression sequences have been manually annotated by a trained workforce (n=23 trained labellers) including medical students and employees of ThinkSono to a) train the algorithm and b) evaluate its performance quantitatively.

### Ultrasound Protocol

Non-enhanced ultrasound imaging was performed by a research physician (at least one year of hands-on ultrasound DVT imaging training) using either Clarius (Clarius L7 (2017) and Clarius L7 HD (2020)) or Philips Lumify L7 or GE VScan Extend (scanned with linear probe) ultrasound devices. Example images for these scanners are shown in Figure 1. Two-point compression ultrasound was used for this study. Clinically, a compression is deemed adequate when the vein was compressed fully or is incompressible at the same pressure a healthy vein would collapse. The femoral vessels were examined from 2 cm distal to the saphenofemoral junction to 2 cm proximal from the inguinal band. The superficial femoral vessels were examined in the adductor canal. The examination of the popliteal vein starts from the distal 2 cm of the popliteal vein and its trifurcation into the anterior tibial vein, posterior tibial vein, and the peroneal vein. The entire examination has been recorded as screen capture and cropped to the ultrasound image area without user-interface content. Participants were positioned in a supine position, with hip rotated outwards by about 60 – 80° and knee flexed at about 60°. The knee area was examined either supine with neutral hip and knee flexed at 80 – 90° or sitting upright with knee hanging loose over the gurney edge at 90°.

**Figure 1:**
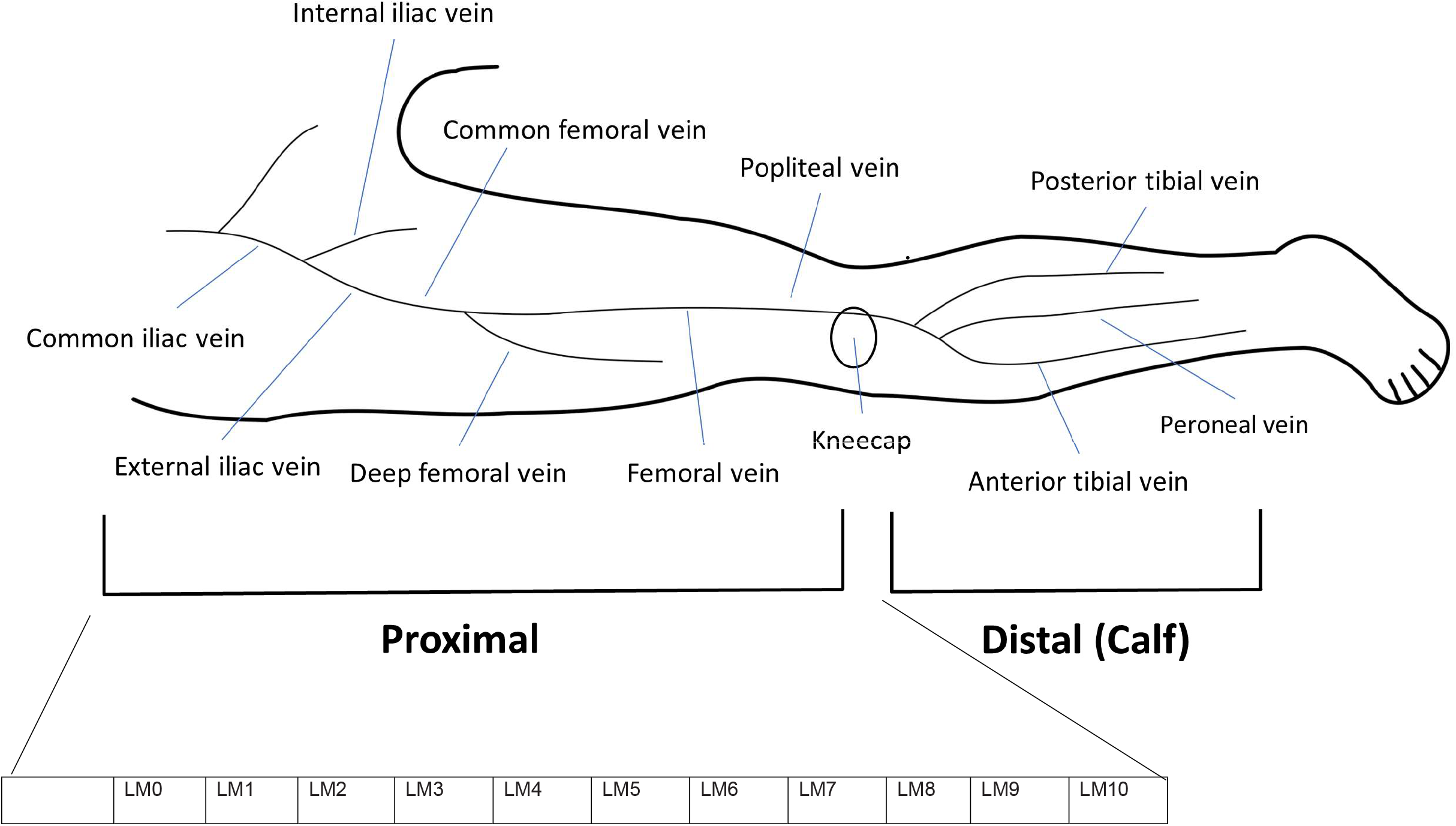
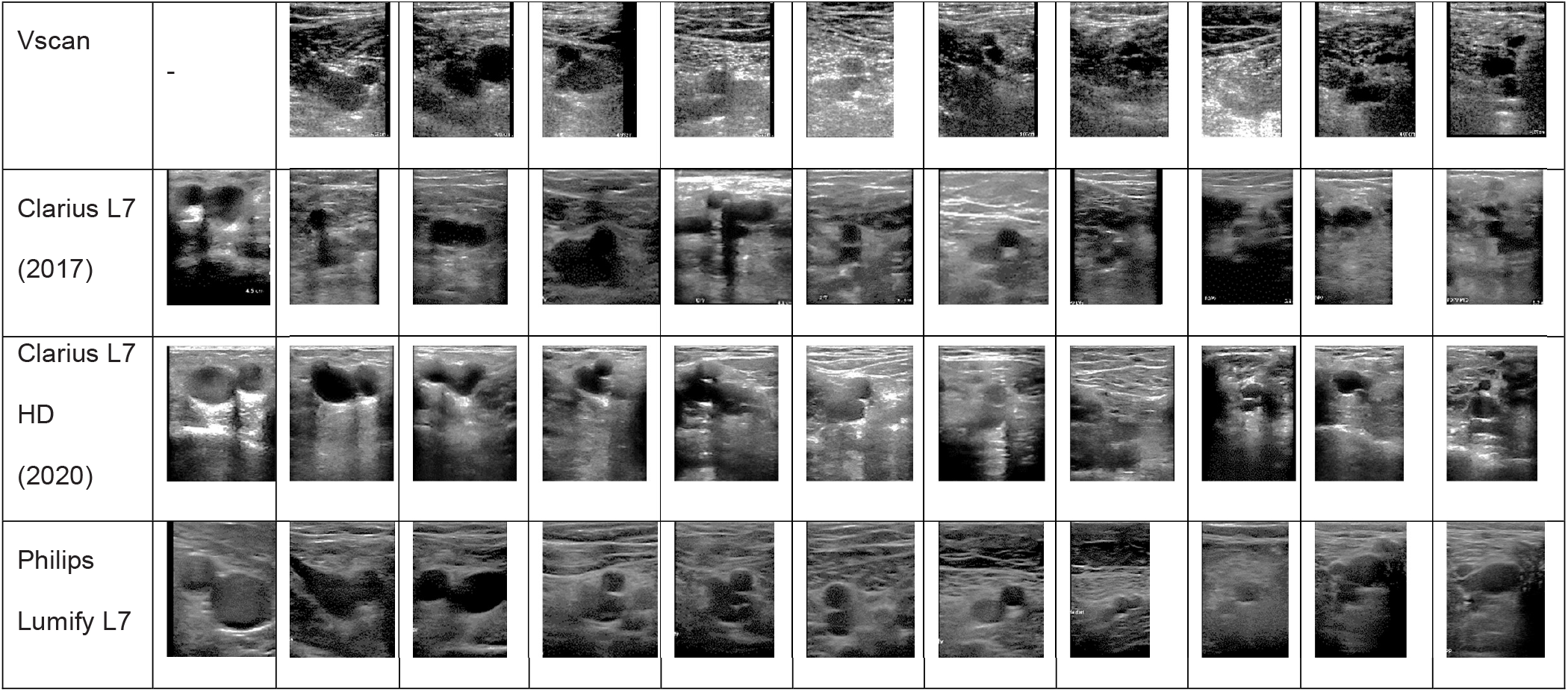
Examples of the chosen anatomically salient landmarks and overview over the investigated anatomy, acquired by different acquisition devices and from different subjects. This figure illustrates the diversity in our dataset. See the overview above the table and Table 1 and 2 in Appendix A for a description for the location of these landmarks. These example images have been manually cropped and contrast normalised for better readability in the manuscript.

### Statistical Analysis

Ultrasound has a specificity of 94% and a sensitivity of 97% for DVT detection^12,13^ when performed by specialised radiologists. Two studies reported sensitivity of 84.4 – 90.0% and specificity of 97.0 – 97.1% when intensely trained nurses and GPs were the ultrasound operators^9,10^.

**Table 1:**
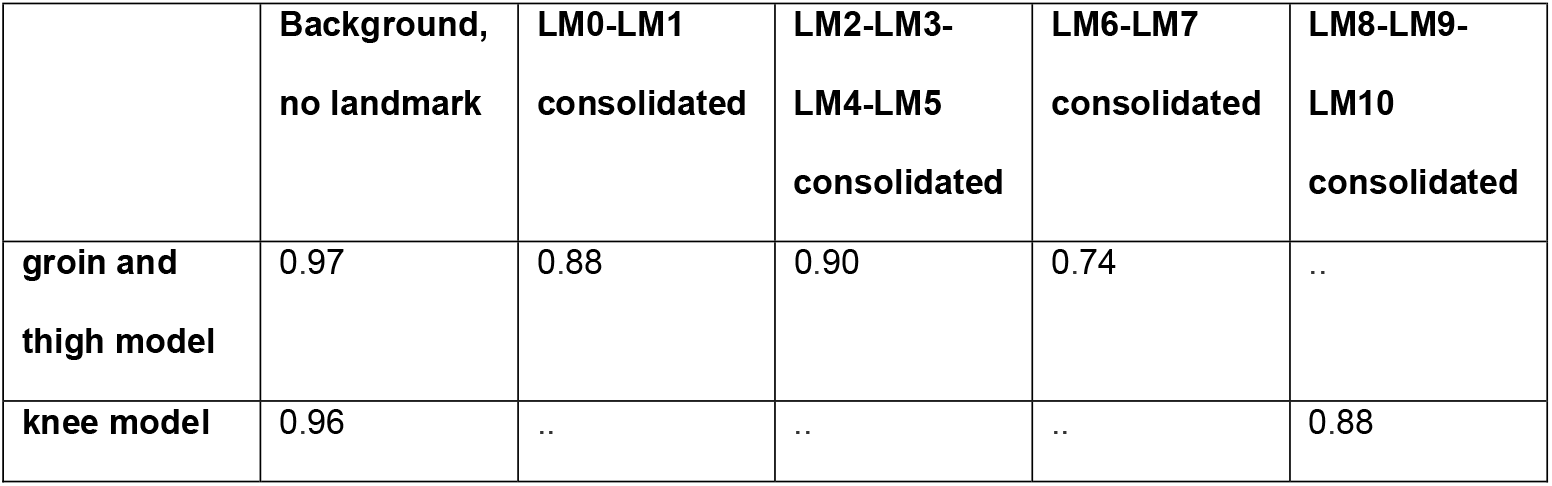
Quantitative results for the landmark detection task of the used models. Evaluation according to Eq. 5 on the internal validation set.

**Table 2:** Quantitative results for the vein compression state task of the used models. Evaluation according to Eq. 5 on the internal validation set.

|  | Vein open | Vein closed and fully<br>compressed |
| --- | --- | --- |
| **groin and thigh model** | 0.92 | 0.95 |
| **knee model** | 0.89 | 0.89 |

### Sample size

The sample size in this manuscript is 167.145 annotated ultrasound imaging frames for model training from 255 healthy volunteers, 15.523 annotated frames from 26 participants for internal model validation and 17.855 frames in video recordings from 51 patients with suspected DVT for external, prospective model evaluation. This is in line with other studies evaluating algorithmic diagnostic decision support, who recently reported external validation sample sizes of, e.g., 50^32,33^, 91^34^, and 198^35^ subjects during retrospective testing and 80^36^to 97^37^during prospective testing.

Statistical approximations for sample size estimation^38^ suggest that eight patients with the required outcome would be sufficient in an external validation set, given an incident rate of 7.1% (Table 4) as observed in our thrombosis clinic. We have included 32 patients with confirmed DVT and 19 patients from the same clinic who were suspected but did not suffer from DVT.

The power of this study is above 0.8 at a significance level of 0.05, with a Cohen’s d effect size of 0.5, when assuming an effect between 0.9 (without software support n^9^=697, n^10^=1,107) and 0.95 (with software support, this study n=51) with a standard deviation of 0.1. For this setting, 51 patients are required as minimum to reach a power of 0.8. The *R* software package (© The R Project for Statistical Computing) has been used for numerical power analysis.

Algorithms are evaluated at the participant level. To evaluate classifier performance, we calculate sensitivity, specificity, positive predictive value, negative predictive value, and overall accuracy for DVT identification for the *internal and external validation sets*.

We also generate the receiver operating characteristic curve of the DVT classification score for the *internal and external validation sets* and calculated the area under the receiver operating characteristic curve (AUC). We show confusion matrices at the optimal algorithm threshold.

### Algorithm design

This study aims to validate the effectiveness of an ML-powered device (AutoDVT) for the diagnosis of proximal DVT. AutoDVT is a CE-marked software product (93/42/EEC 40873) that is coupled to a handheld CE-marked ultrasound machine. The AutoDVT software has two functions: (1) Directing the user to correctly position the ultrasound to complete a thorough scan, (2) Analysing the scan results to confirm the presence/absence of a thrombus.

The software uses a fully automated ML vessel segmentation network with auxiliary branches that predict the anatomical location of the ultrasound image relative to the deep veins in the leg and the compression status of the vein (open or closed). Veins have been labelled by a radiologist to be either open or closed and fully compressed. Two identical networks have been trained: one for the groin/thigh area and the knee area. The subject IDs overlap between training set and internal validation set because a sequence can have multiple landmarks but belong to either a healthy patient or a patient with confirmed DVT. See Table A1 in Appendix A for an overview over the algorithm training data and Table A2 in Appendix A for the internal validation data. Annotations include manual delineations of vein and artery cross sections in the images as well as discrete image-level labels for eleven anatomical locations. To facilitate algorithmic evaluation, we have defined anatomically salient landmarks (LM0 – LM10) on the common femoral vein, superficial femoral vein, and popliteal vein. Example images for these landmarks, acquired with the different ultrasound probes that are used in this study, are shown in Figure 1.

To exclude DVT an operator must follow a protocol as instructed by the software. This protocol mimics the clinical practice of three-point or two-point examinations^14-16^, which means doing compression ultrasound in two to three regions with the greatest risk of developing thrombosis.

For three-point compression protocols, these regions include: (1) the common femoral vein at the level of inguinal crease (LM0 – LM4), (2) the superficial femoral vein superior in the adductor canal (LM5 – LM7), and (3) the popliteal vein and its trifurcation in the popliteal fossa (LM8 – LM10).

For two-point compression protocols the same regions are examined except (2), i.e., LM0 – LM5 in the groin and LM8 – LM10 in the knee. To match the clinical practice in the clinic from where our *external validation set* originates, we investigate the effectiveness of algorithmically evaluated two-point compression DVT examinations in this study.

Thus, using the *training set*, ML models are trained on consolidated groups of landmarks LM0 – LM1, LM2 – LM3 – LM4, i.e., two groups, for (1) and one group, LM8 – LM9 – LM10, for (3). This means three successful vein compressions, two in the groin area and one in the knee area, are required in total to exclude DVT. All identified anatomical locations must show fully compressible veins, otherwise the participant is categorised as suspected DVT case.

Two deep ML networks with identical architecture as shown in Figure 2 were trained on a GPU server (Nvidia Tesla K80) using the Adam optimizer with momentum 0.9 to optimise the parameters of the network. Binary cross entropy (BCE, Eq. 1 and 2) is used for the segmentation task (one-hot encoded) and the vein open/closed task. Cross entropy (CE, Eq. 3) is used for the anatomical landmark detection task as an error metric.

**Figure 2:**
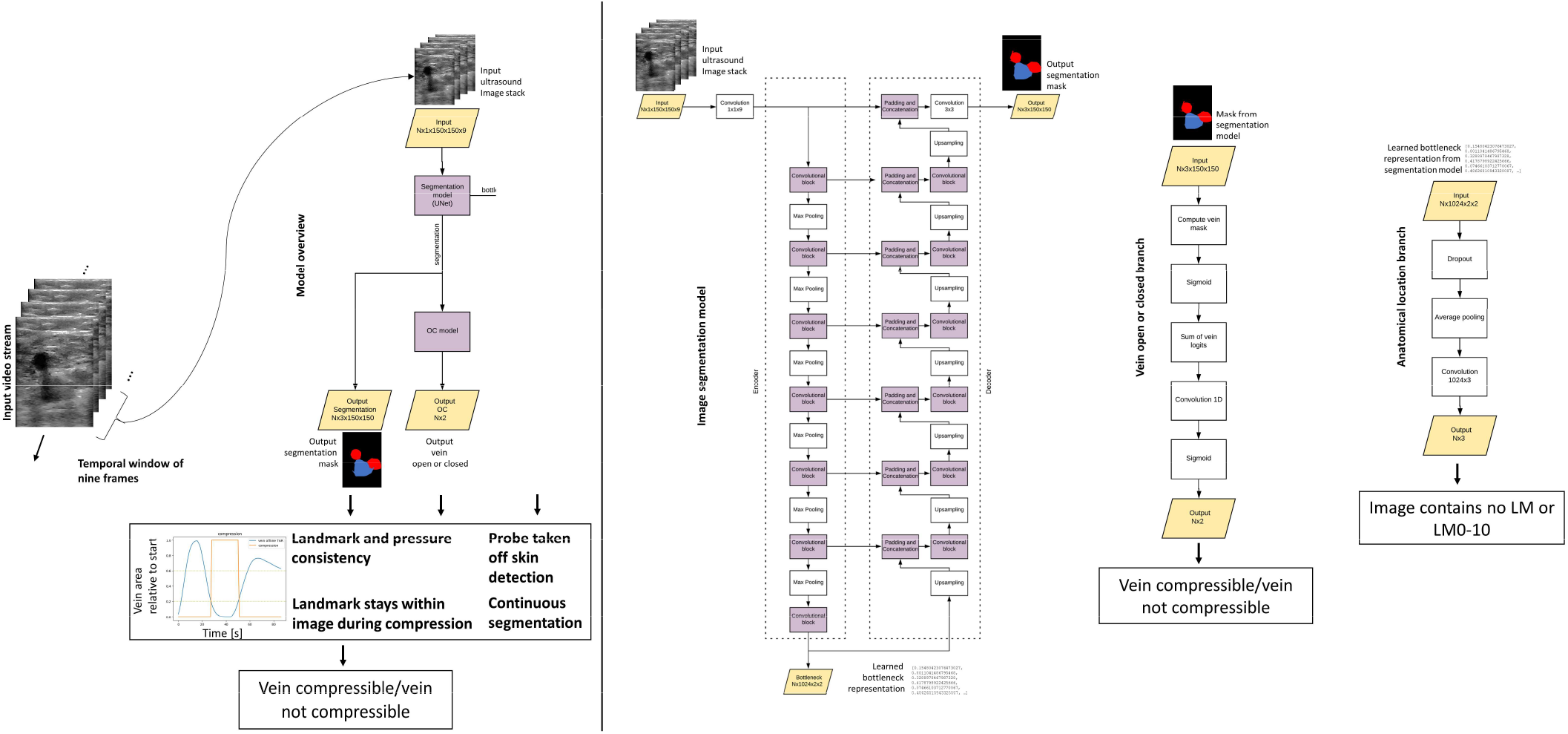
overview over the AutoDVT prototype core algorithm. A U-Net^18^ serves as a backbone for joint vessel masking. The prediction of the anatomical location of the image is based on our previous work^19^. Network branches predict the anatomical location and if the vessel is open or closed under pressure. Landmark predictions are performed from the learned numeric representation in the bottleneck layer; vessel compression state is predicted from the output segmentation mask. The network components are connected and can be trained through backpropagation^20^ in an end-to-end manner. The input is a stack of nine images from an ultrasound video stream that moves by one in a sliding window fashion. A single segmentation mask is produced for the last-most image within approximately 25 ms. Two separate models with identical architecture are trained, one for the groin area (LM0 – LM5) and one for the knee area (LM8– LM10). Each model holds 31 475 527 parameters. (OC = open/close)

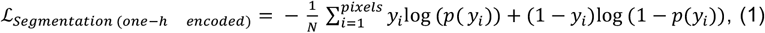

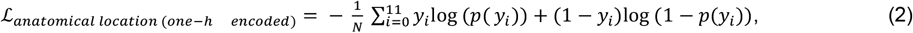

Where *y* is the real label and *p*(*y*) is the predicted probability for the image belonging to this label.

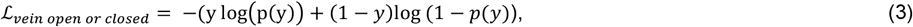

The total error metric (loss function) for our network results as

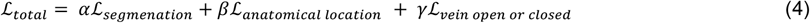

where α and β are adjustable hyper-parameters. We use *α* = 100 *and β* = *γ* = 1.

The pytorch deep learning framework^17^ has been used for implementation.

A series of manually tuned temporal quality control functions ensures robust communication with the user regarding vessel location in the image, quality of compressions, imaging parameters and placement of the probe, see Figure 2.

The *internal validation set* (n=26 healthy subjects, held out from training, Appendix A) has been used to test the models’ performance during development by comparing segmentations to manual delineations of the vessels and manual, categorical image labels with respect to the anatomical locations (LM0 – LM10) and the vessel compression status (open or fully closed).

For categorical labels, the F1-score is used,

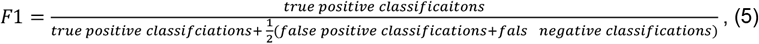

And for segmentation masks the Sørensen–Dice Coefficient is applied per label (background, artery, vein),

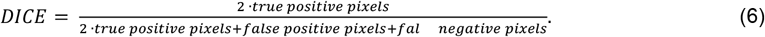

Additionally, the bounding boxes for the individual segmentation masks are generated and the intersection over union (IoU = Jaccard index) is computed, which is a common performance metric for object detection tasks,

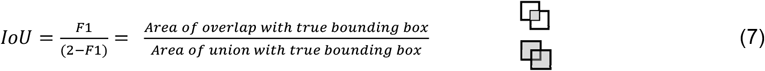

In an end-user scenario an operator would have three attempts to complete a compression, otherwise referral is recommended. A screenshot of the AutoDVT software during use is shown in Figure 3.

**Figure 3:**
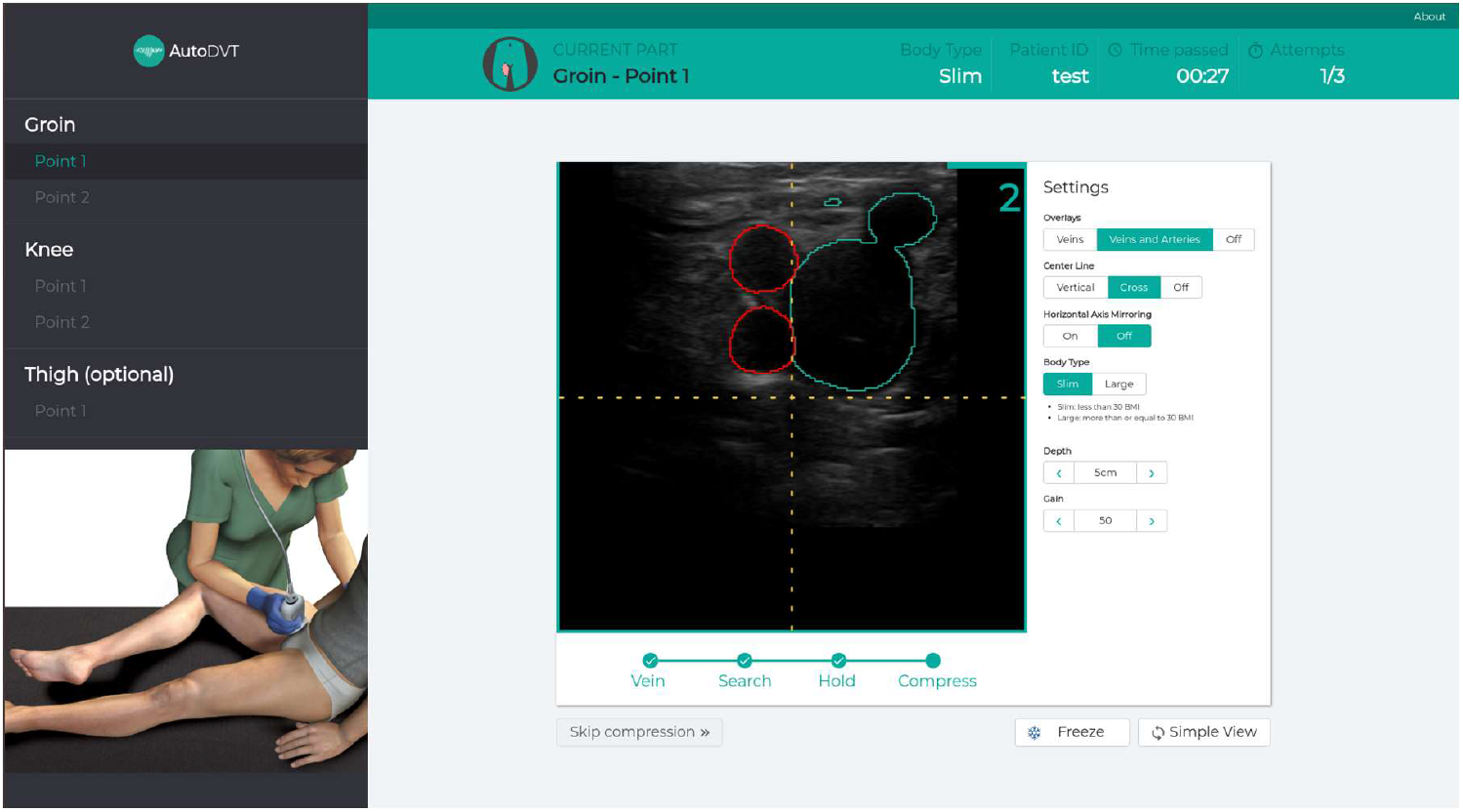
The AutoDVT software instructs users to locate a given landmark, instructs to perform a correct compression and evaluates the result automatically.

### Cost effectiveness

We simulated the potential cost-effectiveness of a ML-enabled approach at the front line of care, where non-specialists may perform the examination independently. A decision tree analytic model was designed and implemented in Microsoft Excel (© Microsoft Corporation) to estimate the lifetime costs and benefit measured in terms of Quality Adjusted Life Years (QALYs) for different proximal deep vein thrombosis (DVT) testing algorithms. The current clinically used diagnostic DVT algorithm is shown in Figure 4a and the proposed integration of our method is shown in Figure 4b.

**Figure 4:**
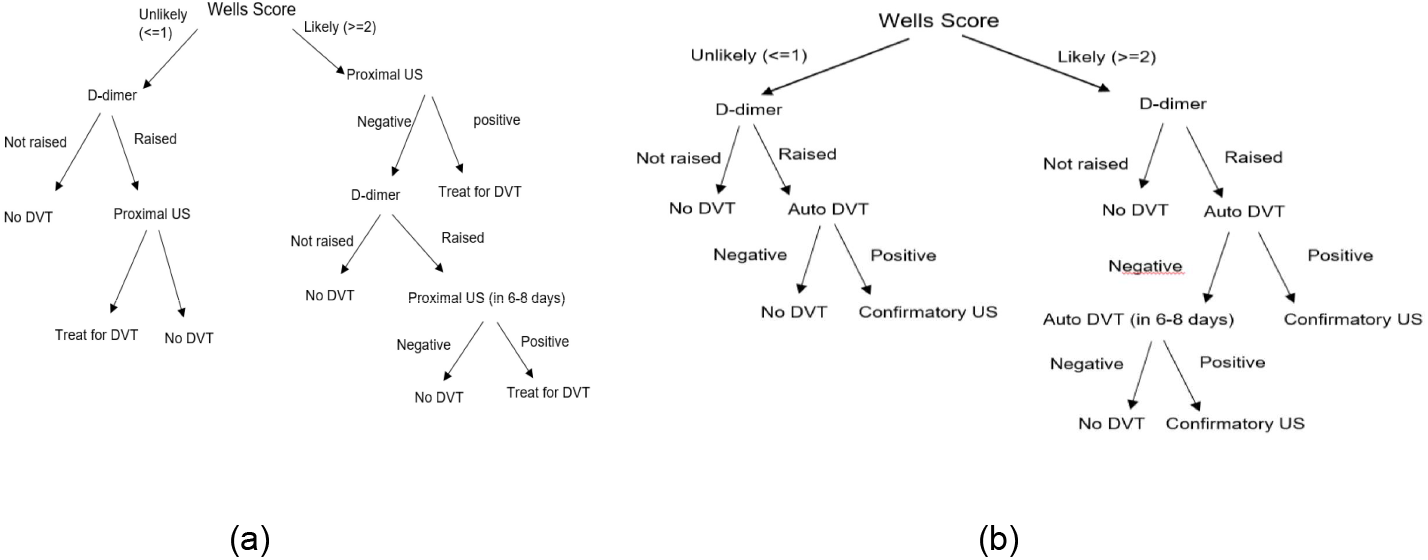
(a) current clinical algorithm to diagnose DVT and (b) the proposed modification with our approach to support front line of care staff to perform the diagnosis reliably.

The cost analysis model adheres to guidelines issued by the National Institute of Health and Care Excellence (NICE)^21^. It uses an NHS and personal social services perspective with costs at 2018/19 prices and with discounting for both costs and QALYs being undertaken at 3.5% per annum.

The model uses sensitivity (the ability of a test to correctly identify a patient with a true proximal DVT) and of specificity (the ability of a test to correctly identify a patient without a true proximal DVT) as measured on the *external validation set* in this study. We also include clinical tests (Wells Score, D-dimer, and proximal ultrasound) that form part of the diagnostic algorithm.

The cost analysis model splits patients into two subgroups at the start of each algorithm, a subgroup in which patients have a proximal DVT and a subgroup in which patients do not have a proximal DVT. Measured sensitivity and specificity values are used alongside an estimate of the prevalence of proximal DVT of 14.7% taken from Kilroy et al.^22^ to estimate the number of patients (from a cohort of user specified size) that receive each clinical test and their ultimate diagnoses (proximal DVT or not). Patients with a diagnosed proximal DVT will receive treatment.

The cost analysis model generates four possible outcomes for patients based on their DVT status and the results of each diagnostic algorithm: Treated patients with a true DVT (true positive patients), treated patients without a true DVT (false positive patients), untreated patients without a true DVT (true negative patients) and untreated patients with a true DVT (false negative patients).

Each of the four diagnostic accuracy outcomes have estimated associated costs incurred and utility accrued for the patients. These numbers are multiplied by the proportion of patients in each outcome and are combined with the costs of each test to obtain estimates of the total costs and QALYs for the diagnostic algorithm. When costs and QALYs are obtained for a diagnostic algorithm including the ML model and one excluding our approach, the estimated incremental cost-effectiveness ratio for AutoDVT can be calculated. Details regarding model parameters are provided in Appendix B.

### Data availability

Beginning 9 months and ending 36 months following article publication in a peer reviewed journal algorithm raw data results access in line with the informed consent of the participants, subject to approval by the project ethics board and under a formal Data Sharing Agreement. Proposals may be submitted up to 36 months following article publication in a peer reviewed journal. Custom computer code is available through ThinkSono Ldt with case specific license agreements.

## Results

### Study participation

124 patients who presented to the DVT diagnostic clinic with symptoms suggestive of DVT were approached for inclusion into this study, thus, for inclusion into the *external validation set*.

36 patients have been excluded during the enrolment phase for various reasons as summarised in the Consort Diagram in Figure 5. Two patients with confirmed DVT have been excluded due to imaging conditions that are not covered by the standard compression ultrasound DVT protocol (non-echogenic thrombus and superior thrombosis in the iliac vein). Control participants had no DVT based on comprehensive clinical and laboratory testing performed under the supervision of and interpreted by a haematologist. This results in a data set comprising of 88 eligible patients. An overview of patient characteristics in this clinic’s database is given in Table 4.

**Figure 5:**
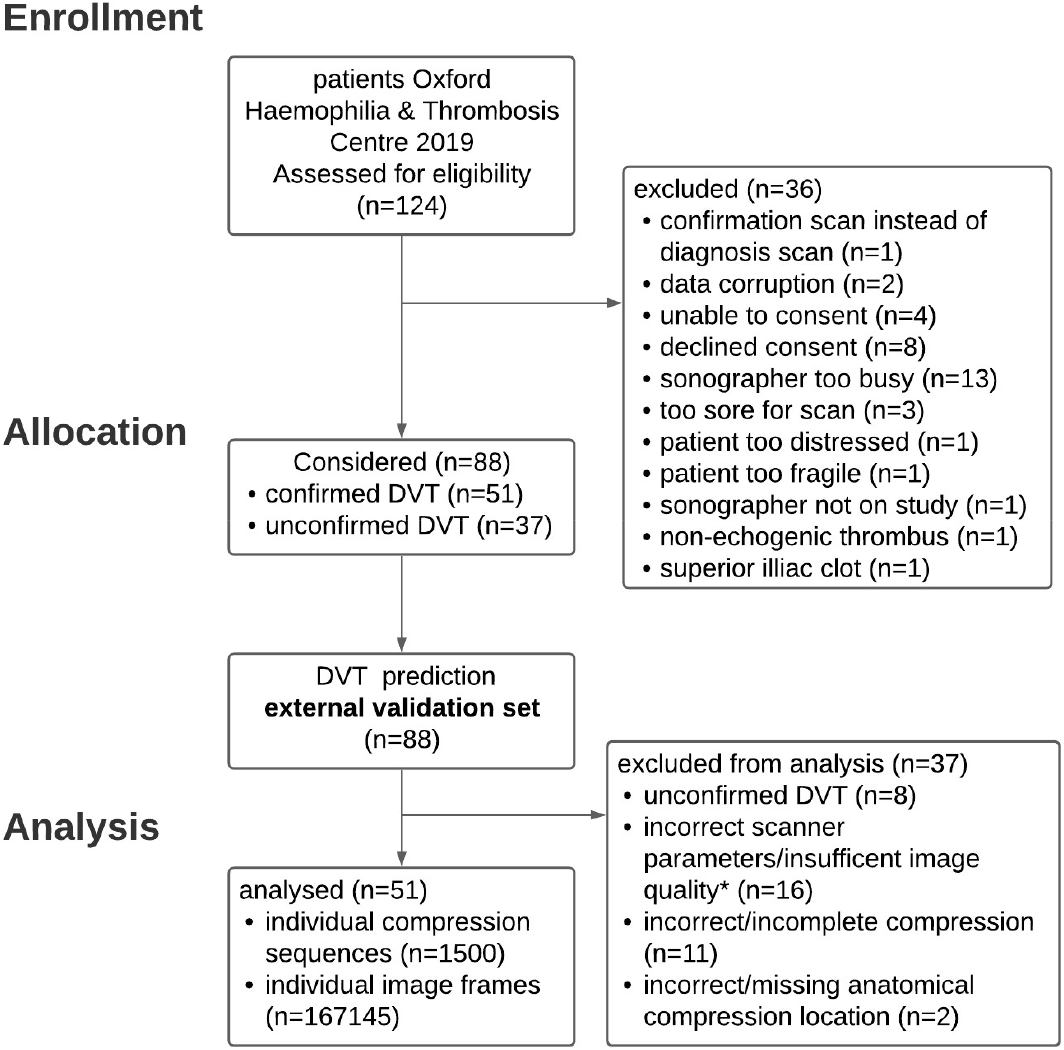
Consort diagram for study enrolment, allocation, and analysis.

It was specified that all examinations that were not performed according to the standard implemented in our study design should be omitted, thus, secondary exclusion criteria must be applied. Hence, 37 patients (19 DVT positive, 18 DVT negative) have been further excluded during the analysis phase, due to, radiologist/haematologist confirmed, incorrect/incomplete compression (11), compression on incorrect/missing anatomical location (2), incorrect scanner parameters evaluated by 10-point expert image quality scoring (16). After these exclusions, the remaining sequences of a positive DVT patient may not include the clip that shows the positive DVT. Thus, further eight sequences belonging to a positive DVT case with a missing compression sequence confirming the DVT have been excluded. Of the remaining 51 patients 32 patients are DVT positive and 19 DVT negative, confirmed by to the current clinical pathway. This results in 100 individual compression sequences on defined anatomical vessel locations in these patients.

### Participant Characteristics of the *external validation set*

The external validation set was drawn from the general population of 2041 patients with suspected DVT from the Haemophilia and Thrombosis Centre at University of Oxford. The characteristics of the entire population during the year 2019 is summarised in Table 4. The ethical approval in place did not allow for the collection of these characteristics for the individual patients that have been enrolled into this study.

### Algorithm performance on the *internal validation set*

Figure 6 shows qualitative examples for the segmentation output of our method. Table 1 shows quantitative results for the anatomical landmark detection task; Table 2 for the vessel compression task and Table 3 regarding segmentation performance. Common image evaluation metrics, Sørensen–Dice Coefficient (Eq. 6) for segmentation results and F1-score (Eq. 5) for anatomical landmark discrimination and categorical vessel compression analysis, are used for quantitative evaluation.

**Table 3:** Quantitative results for the vessel segmentation task of the used models. Evaluation according to Eq. 6 and Eq. 7 on the internal validation set.

|  | Sørensen–Dice Coefficient |  |  | Bounding-box<br>intersection over<br>union |
| --- | --- | --- | --- | --- |
|  | background | artery | vein |  |
| **groin and thigh model** | 0.99 | 0.89 | 0.75 | 0.70 |
| **knee model** | 0.99 | 0.81 | 0.88 | 0.85 |
| **metric** | Eq. 6 |  |  | Eq. 7 |

**Figure 6:**
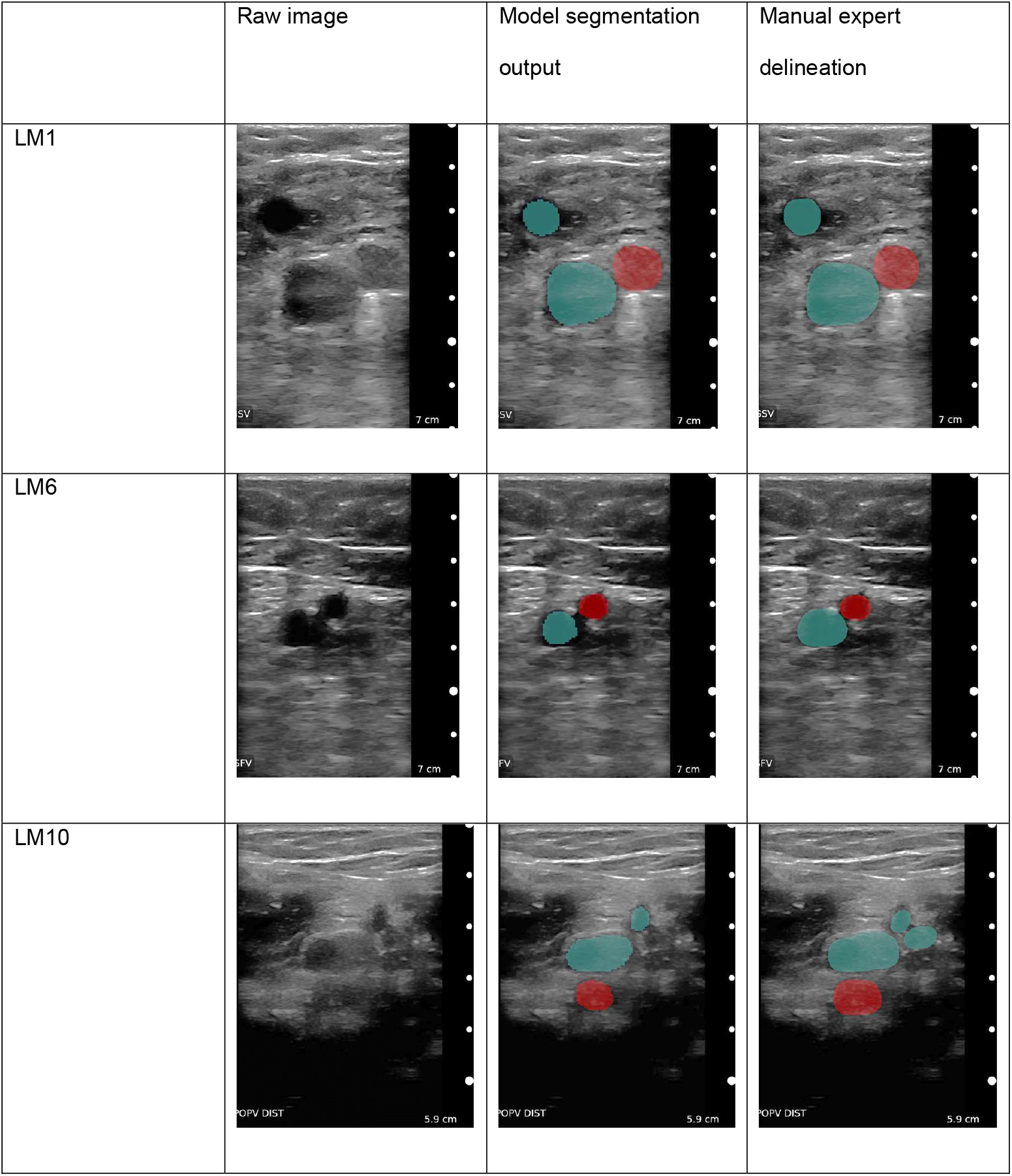
Qualitative example images for our model’s segmentation performance. The segmentation is robust throughout compressions. The vein area is evaluated for complete compressibility to exclude DVT. Device: Clarius L7 (2017).

### Algorithm performance on the *external validation set*

The results of the analysis of the enrolled patients is summarised in Table 5. Receiver operator curves are shown together with the confusion Matrix in Figure 7 on patient level and Figure 8 on sequence/anatomical landmark level. Note that these results are based on retrospective analysis of prospectively acquired ultrasound videos. In a fully prospective setting, AutoDVT guides the operator to acquire the correct images and to perform correct compressions. This implicitly reflects the data curation we performed on the external validation set and the performance is expected to be similar when using our method for diagnosis.

**Table 4:**
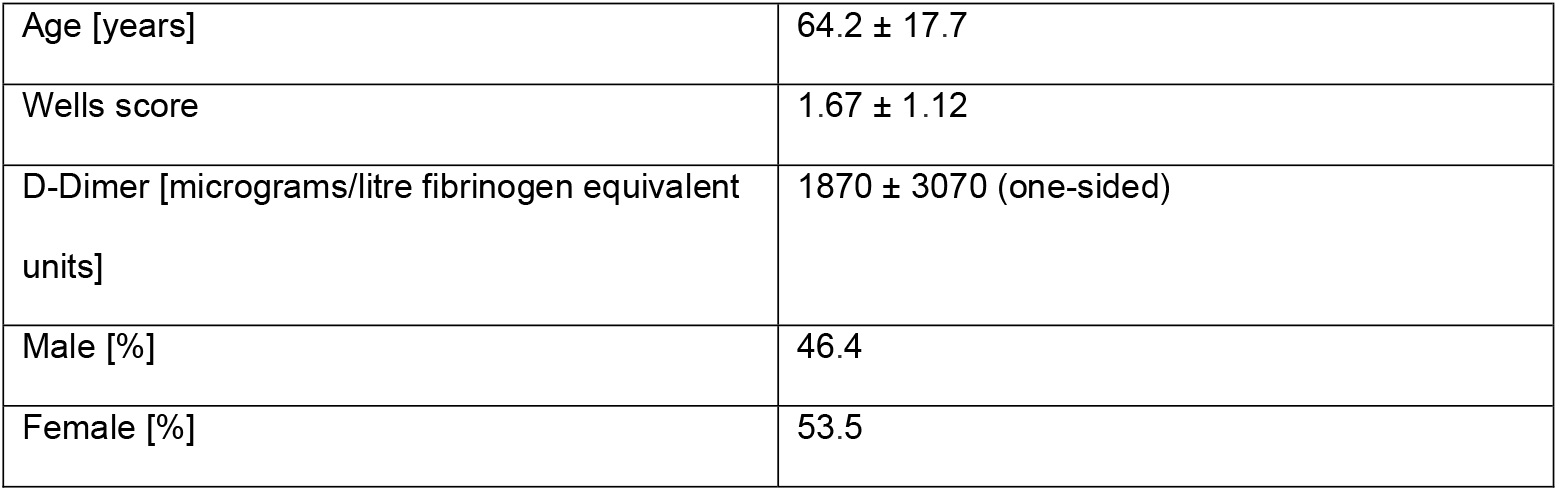
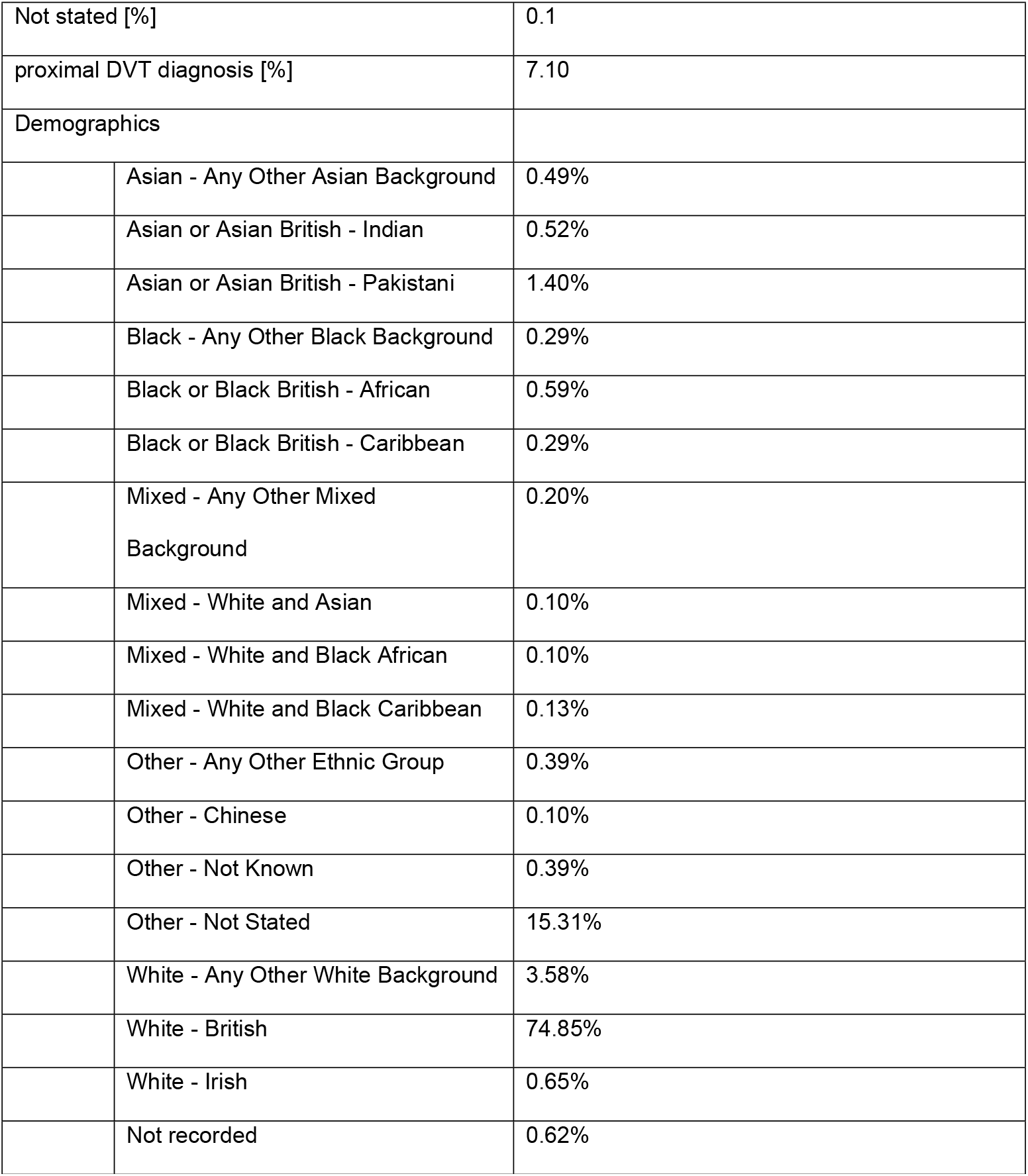
general population overview for model training and external validation set.

**Table 5:** Values are expressed between [0,1] intervals. NPV = negative predictive value, PPV = positive predictive value.

| Performance metrics | Algorithm decision |
| --- | --- |
| Sensitivity | **0.938** |
| Specificity | **0.842** |
| PPV | **0.909** |
| NPV | **0.889** |
| Accuracy | **0.902** |

**Figure 7:**
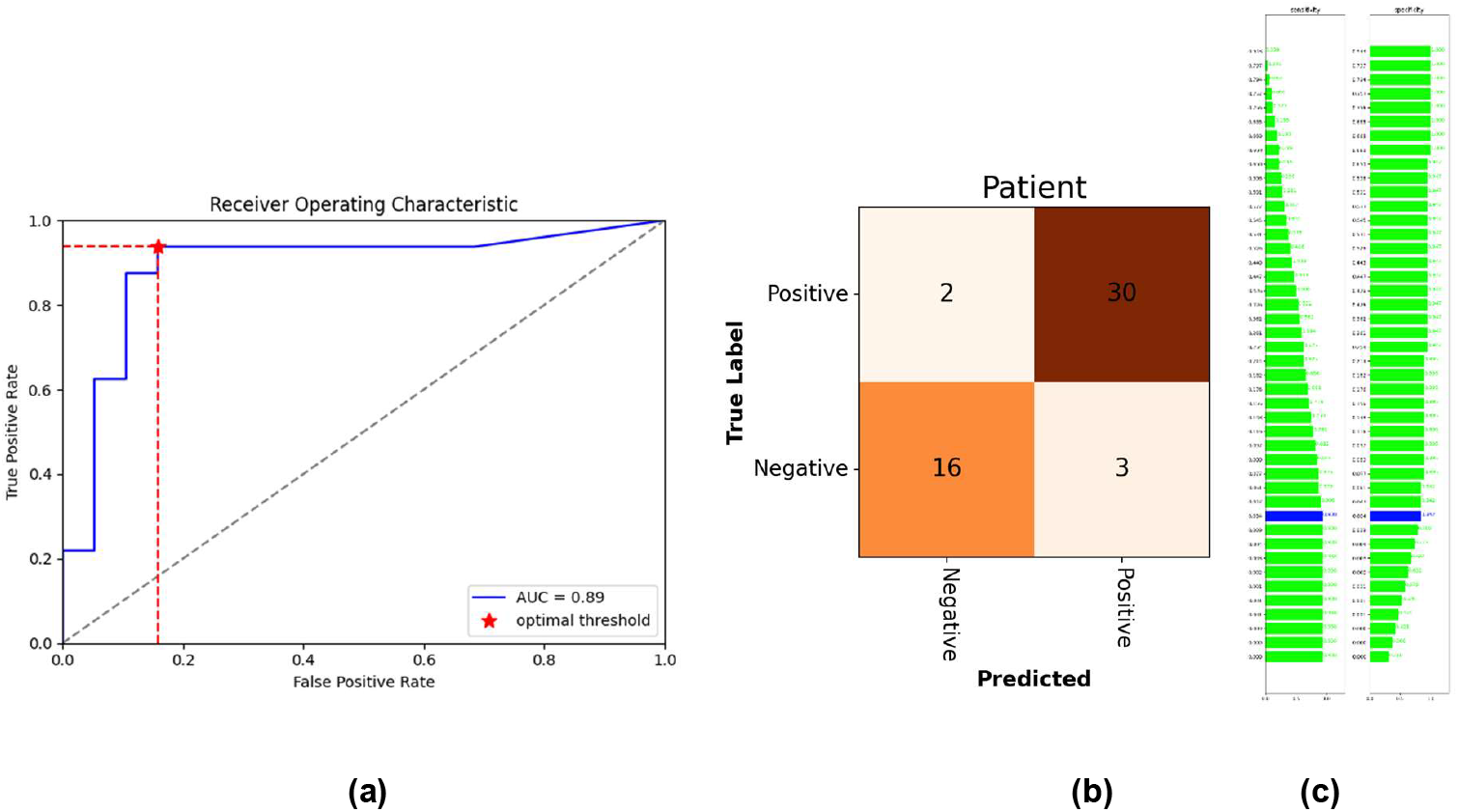
Receiver operator characteristics for the final examination algorithm. Confusion matrix (b) at optimal threshold (c). Vessel status is extracted automatically through the ML models from 51 enrolled patients.

**Figure 8:**
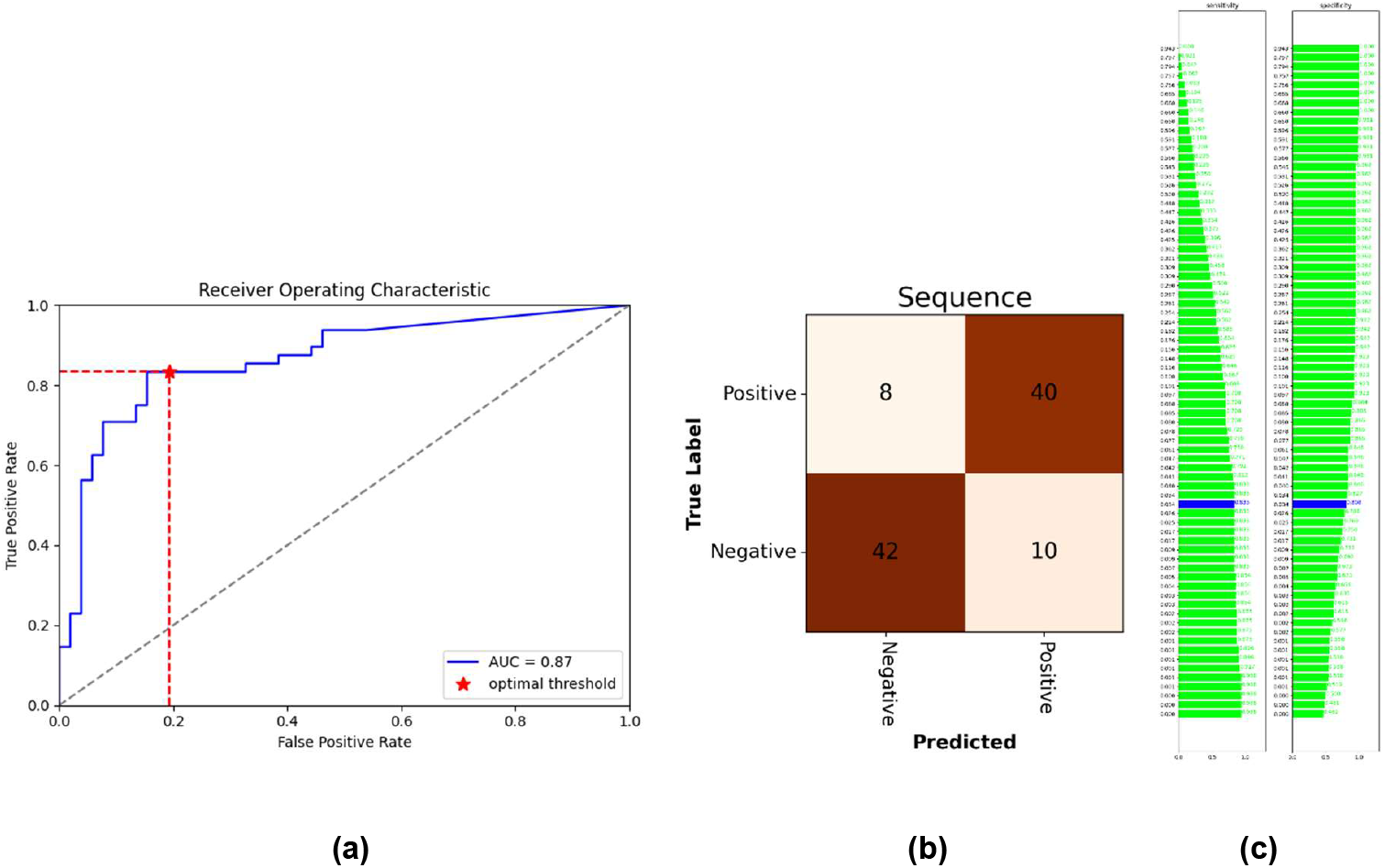
Receiver operator characteristics for the correct compression classification per ultrasound sequence/anatomical landmark. Confusion matrix (b) at optimal threshold (c). Vessel status is extracted automatically through the ML models from the 100 available anatomical landmark sequences.

### Cost effectiveness

With the sensitivity and the specificity from this study a net monetary benefit (NMB) of up to $150 per ML-supported examination can be achieved using a willingness to pay of $26 000 per QALY^23^. Figure 9 shows how the NMB changes with different prices for such an examination.

**Figure 9:**
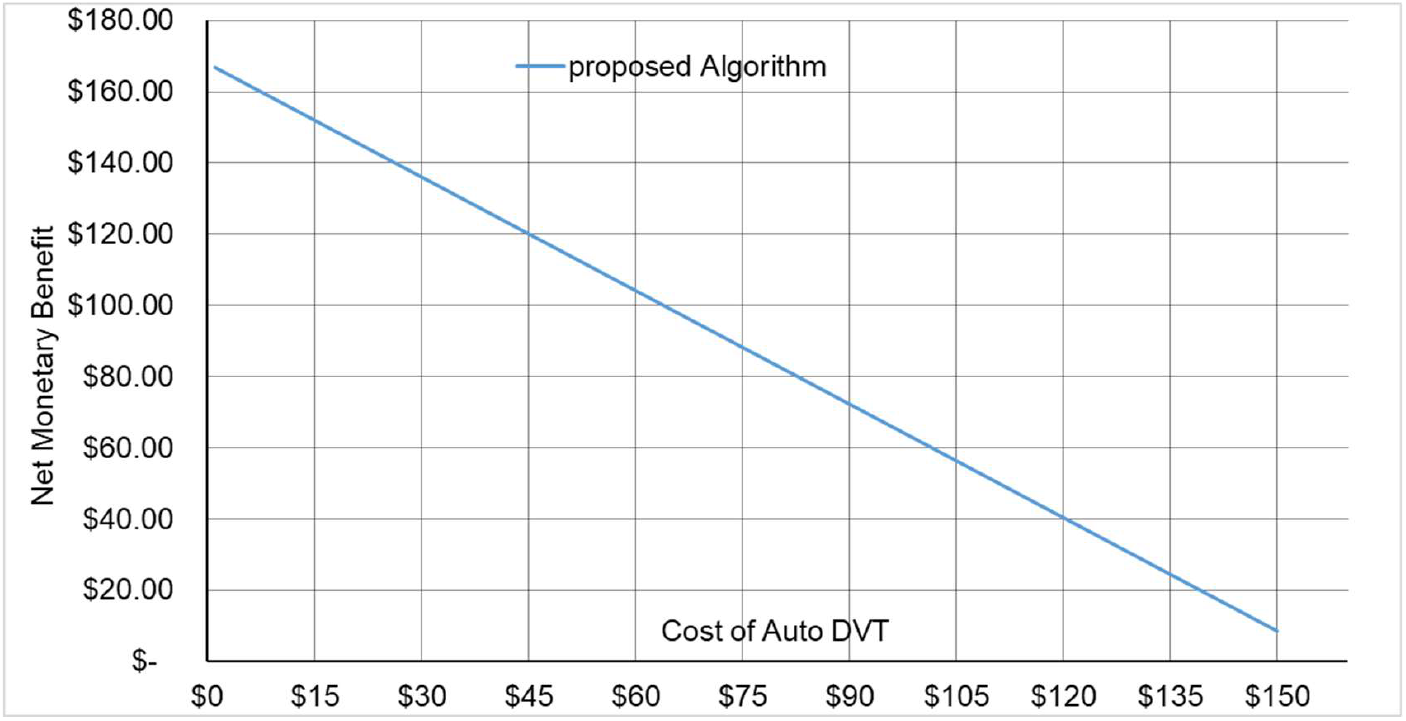
Costs of the guidance tool vs. net monetary benefit per examination when implementing ML guided DVT diagnostics into a clinical algorithm as it is shown in Figure 4b.

## Discussion

This study provides a proof of concept that ML-based analysis can accurately distinguish patients with and without DVT while providing guidance for image acquisition according to the clinical standard. Evaluation was performed on a sample size of n=51 enrolled patients from the same clinic, 32 DVT-positive patients and 19 DVT-negative patients. Algorithmic DVT diagnosis results in a sensitivity of 93.8% and a specificity of 84.2%, a positive predictive value of 90.9%, and a negative predictive value of 88.9%. Furthermore, a cost analysis simulation model has been evaluated when integrating the proposed algorithm into the clinical practice. Assuming a willingness to pay threshold of $26 000/QALY^23^, a net monetary benefit of up to $150 per examination could be attained when ML guidance is used by the front line of care for DVT diagnosis.

Although ML has been studied for a variety of diagnostic approaches^24-26^, to the best of our knowledge this is the first study that has shown potential benefits for the diagnosis of DVT. The dominating application area of ML is the automated analysis of static images (CT, MRI, etc.). Free-hand ultrasound poses additional challenges compared to these approaches.

First, a user needs to be directed and guided to acquire images, which are suitable to make a prediction though a ML model. This requires algorithmic provisions to discriminate useful images from images that do not adhere to a clinic standard. We solved this problem through training a discriminator ML model, which can identify predefined anatomical locations along the femoral vein.

Second, compression ultrasound requires the analysis of continuous image sequences which is challenging in a setup that requires real-time feedback. We solve this problem through a sliding window, multi-channel input approach, which enforces spatio-temporal consistency for a combined vein-segmentation with learned decision boundaries for identifying a vessel as fully closed.

Furthermore, mobile ultrasound probes are used and connected to a GPU-accelerated laptop to provide sufficient computational power.

Third, image domain shift is a serious limitation of ML applications in Healthcare. Domain shift occurs when a model is trained on images that have been acquired on one device while the testing is performed on images from other, previously unseen images from different devices. Commonly, a noticeable drop in performance is observed in such situations. We mitigate this problem through integrating image data from a diverse set of devices, covering almost the entire market for mobile ultrasound devices. Still, there is no established method for robust domain adaptation^27^. Hence, a risk of reduced performance remains when applying the presented algorithms to images from a new device. This risk must be avoided by deploying these algorithms exclusively with thoroughly tested, specific devices.

ML-supported devices such as described here are often summarised as clinical AI^26^. A critical element of any AI-based support tool is its clinical relevance.

The major strengths of this study are the development and validation of an algorithm and system that has the potential to push DVT screening upstream in clinical pathways and to enable front line of care personnel to acquire data at the standard of an expert. We provide a possible integration strategy into a diagnostic DVT decision tree and show that the use of AI technology can be cost effective. We would expect that rapid point of care diagnostics and wide availability of testing, which is enabled by our approach, would lead to timely treatment, decreased stress, and increased patient satisfaction.

Our study has several limitations. First, we evaluated a prospectively enrolled patient cohort retrospectively, on video sequences that have not necessarily been acquired at an optimal standard. Therefore, we had to curate the data and automatically extract clips from entire exam video recordings that would be most similar to clips as they would be acquired by the discussed software guidance method. Furthermore, free-hand ultrasound examinations are highly operator-dependent, and every operator has a unique style of examination. Our proposed approach aims to standardise these styles to provide optimal input for subsequent image analysis parts and to aid clinical audits.

Second, our patient cohort is small, and we compare across-population with findings from literature. This limits the types of statistical techniques that can be employed in this study to evaluate statistical significance. We are currently conducting a multi-centre prospective trial which will address these issues to give further insights into the practical implications of employing AI support for DVT diagnosis.

In conclusion, our study shows the potential of a ML-powered system using free-hand ultrasound to identify DVT in clinical populations with high-throughput requirements and at primary care level. Since access to ultrasound imaging is increasing, especially mobile ultrasound devices, which can currently be purchased for $3 000-$9 000, a ML-supported examination by less specialised front line care workers has the potential to be adopted for proximal DVT screening before confirmatory tests.

## Author contributions

B.K., N.C., A.M., M.H., M.D.S. conceived and planned the experiments; B.K., R.M. and F.AN. conducted literature research; A.M. designed the model and the computational framework and analysed the data; S.M. and F.AN. carried out the implementation; B.K. wrote the manuscript with input from all authors; A.M., A.C.R., and M.D.S. designed and performed the experiments; J.O. and R.M. assisted with anatomical measurements; B.K. and N.C. devised the project, the main conceptual ideas and proof outline. A.M., J.O., S.M., and F.AN. verified the data; Ch.D. managed patient enrolment, patient consent and patient characteristics. N.C., M.H., R.M., A.C.R., M.D.S. and B.K. revised the final manuscript. All authors discussed the results and contributed to the final manuscript.

## References

1. ISTH Steering Committee for World Thrombosis Day. Thrombosis: a major contributor to the global disease burden. J Thromb Haemost. 2014; 12: 1580 – 1590.

2. Jha AK, Larizgoitia I, Audera-Lopez C, Prasopa-Plaizier N, Waters H, Bates DW. The global burden of unsafe medical care: analytic modelling of observational studies. BMJ quality & safety. 2013 Oct 1;22(10):809–15.

3. Cohen AT, Agnelli G, Anderson FA, Arcelus JI, Bergqvist D, Brecht JG, Greer IA, Heit JA, Hutchinson JL, Kakkar AK, Mottier D, Oger E, Samama MM, Spannagl M; VTE Impact Assessment Group in Europe (VITAE). Venous thromboembolism (VTE) in Europe. The number of VTE events and associated morbidity and mortality. Thromb Haemost. 2007; 98(4): 756–64.

4. Beckman MG, Hooper WC, Critchley SE, Ortel TL. Venous thromboembolism: a public health concern. American journal of preventive medicine. 2010 Apr 1;38(4):S495–501.

5. Plüddemann A, Thompson M, Price CP, Wolstenholme J, Heneghan C. The D-Dimer test in combination with a decision rule for ruling out deep vein thrombosis in primary care: diagnostic technology update. British Journal of General Practice. 2012 May 1;62(598):e393–5.

6. NICE Guidance. [Online].; 2019. Available from: https://www.nice.org.uk/guidance/cg144/chapter/recommendations.

7. Elliott CG, Goldhaber SZ, Jensen RL. Delays in diagnosis of deep vein thrombosis and pulmonary embolism. Chest. 2005 Nov 1;128(5):3372–6.

8. Storck M, Bauersachs R. Venenthrombose und Lungenembolie. In Chirurgie Basisweiterbildung 2013 (pp. 452–458). Springer, Berlin, Heidelberg. AWMF Leitlinien-Register N0. Venenthrombose und Lungenembolie: Diagnostik und Therapie. [Online].; 2015. Available from: https://www.awmf.org/leitlinien/detail/ll/065-002.html.

9. Mumoli N, Vitale J, Cocciolo M, Cei M, Brondi B, Basile V, Sabatini S, Gambaccini L, Carrara I, Camaiti A, Giuntoli S. Accuracy of nurse-performed compression ultrasonography in the diagnosis of proximal symptomatic deep vein thrombosis: a prospective cohort study. Journal of Thrombosis and Haemostasis. 2014 Apr;12(4):430–5.

10. Mumoli N, Vitale J, Giorgi-Pierfranceschi M, Sabatini S, Tulino R, Cei M, Bucherini E, Bova C, Mastroiacovo D, Camaiti A, Palmiero G. General practitioner–performed compression ultrasonography for diagnosis of deep vein thrombosis of the leg: a multicenter, prospective cohort study. The Annals of Family Medicine. 2017 Nov 1;15(6):535–9.

11. Fox JC, Bertoglio KC. Emergency physician performed ultrasound for DVT evaluation. Thrombosis. 2011;2011.

12. Goodacre S, Sampson F, Thomas S, van Beek E, Sutton A. Systematic review and meta-analysis of the diagnostic accuracy of ultrasonography for deep vein thrombosis. BMC medical imaging. 2005 Dec 1;5(1):6.

13. Zierler BK. Ultrasonography and diagnosis of venous thromboembolism. Circulation. 2004 Mar 30;109(12_suppl_1):I–9.

14. Lee JH, Lee SH, Yun SJ. Comparison of 2-point and 3-point point-of-care ultrasound techniques for deep vein thrombosis at the emergency department: a meta-analysis. Medicine. 2019 May;98(22).

15. Zuker-Herman R, Dangur IA, Berant R, Sitt EC, Baskin L, Shaya Y, Shiber S. Comparison between two-point and three-point compression ultrasound for the diagnosis of deep vein thrombosis. Journal of thrombosis and thrombolysis. 2018 Jan 1;45(1):99–105.

16. Dehbozorgi A, Damghani F, Mousavi-Roknabadi RS, Sharifi M, Sajjadi SM, Hosseini-Marvast SR. Accuracy of three-point compression ultrasound for the diagnosis of proximal deep-vein thrombosis in emergency department. Journal of Research in Medical Sciences: The Official Journal of Isfahan University of Medical Sciences. 2019;24.

17. Paszke A, Gross S, Massa F, Lerer A, Bradbury J, Chanan G, Killeen T, Lin Z, Gimelshein N, Antiga L, Desmaison A. Pytorch: An imperative style, high-performance deep learning library. InAdvances in neural information processing systems 2019 (pp. 8026–8037).

18. Ronneberger O, Fischer P, Brox T. U-net: Convolutional networks for biomedical image segmentation. InInternational Conference on Medical image computing and computer-assisted intervention 2015 Oct 5 (pp. 234–241). Springer, Cham.

19. Tanno R, Makropoulos A, Arslan S, Oktay O, Mischkewitz S, Al-Noor F, Oppenheimer J, Mandegaran R, Kainz B, Heinrich MP. Autodvt: Joint real-time classification for vein compressibility analysis in deep vein thrombosis ultrasound diagnostics. InInternational Conference on Medical Image Computing and Computer-Assisted Intervention 2018 Sep 16 (pp. 905–912). Springer, Cham.

20. LeCun Y, Boser B, Denker JS, Henderson D, Howard RE, Hubbard W, Jackel LD. Backpropagation applied to handwritten zip code recognition. Neural computation. 1989 Dec;1(4):541–51.

21. National Institute of Health and Care Excellence L. https://www.nice.org.uk/process/pmg9/resources/guide-to-the-methods-of-technology-appraisal-2013-pdf-2007975843781. [Online].; 2013.

22. Kilroy DA, Ireland S, Reid P, Goodacre S, Morris F. Emergency department investigation of deep vein thrombosis. Emergency medicine journal. 2003 Jan 1;20(1):29–32.

23. Goodacre S, Stevenson M, Wailoo A, Sampson F, Sutton AJ, Thomas S. How should we diagnose suspected deep-vein thrombosis?. Journal of the Association of Physicians. 2006 Jun 1;99(6):377-88.

24. De Fauw J, Ledsam JR, Romera-Paredes B, Nikolov S, Tomasev N, Blackwell S, Askham H, Glorot X, O’Donoghue B, Visentin D, van den Driessche G. Clinically applicable deep learning for diagnosis and referral in retinal disease. Nature medicine. 2018 Sep;24(9):1342–50.

25. Bai W, Sinclair M, Tarroni G, Oktay O, Rajchl M, Vaillant G, Lee AM, Aung N, Lukaschuk E, Sanghvi MM, Zemrak F. Automated cardiovascular magnetic resonance image analysis with fully convolutional networks. Journal of Cardiovascular Magnetic Resonance. 2018 Dec 1;20(1):65.

26. Akkus Z, Cai J, Boonrod A, Zeinoddini A, Weston AD, Philbrick KA, Erickson BJ. A Survey of Deep-Learning Applications in Ultrasound: Artificial Intelligence–Powered Ultrasound for Improving Clinical Workflow. Journal of the American College of Radiology. 2019 Sep 1;16(9):1318–28.

27. Wang M, Deng W. Deep visual domain adaptation: A survey. Neurocomputing. 2018 Oct 27;312:135–53.

28. Comissioning OC. Version 2.22018-19 update September Occg Service Specification (2018/19) Primary Care Service Fordvt Testing (updated September 18). 2018 Sept. [Online]. 2018; Available from: https://www.oxfordshireccg.nhs.uk/professional-resources/documents/primary-care/locally-commissioned-services-2018%20-2019/dvt-testing.pdf

29. The Department of Health 2. London: Department of Health. [Online].; 2020]. Available from: https://www.gov.uk/government/publications/nhs-reference-costs-2015-to-2016.

30. Curtis LA, BA. Canterbury: Personal Social services Research Unit. [Online].; 2020. Available from: https://www.pssru.ac.uk/project-pages/unit-costs/unit-costs-2018/.

31. Royal Pharmaceutical Society. British National Formulary. [Online].; 2020. Available from: https://www.medicinescomplete.com/#/browse/bnf.

32. Patel BN, Rosenberg L, Willcox G, Baltaxe D, Lyons M, Irvin J, Rajpurkar P, Amrhein T, Gupta R, Halabi S, Langlotz C. Human–machine partnership with artificial intelligence for chest radiograph diagnosis. NPJ digital medicine. 2019 Nov 18;2(1):1–0.

33. Kanber B, Nachev P, Barkhof F, Calvi A, Cardoso J, Cortese R, Prados F, Sudre CH, Tur C, Ourselin S, Ciccarelli O. High-dimensional detection of imaging response to treatment in multiple sclerosis. NPJ digital medicine. 2019 Jun 10;2(1):1–0.

34. Miura K, Goto S, Katsumata Y, Ikura H, Shiraishi Y, Sato K, Fukuda K. Feasibility of the deep learning method for estimating the ventilatory threshold with electrocardiography data. NPJ digital medicine. 2020 Oct 29;3(1):1–7.

35. Huang SC, Kothari T, Banerjee I, Chute C, Ball RL, Borus N, Huang A, Patel BN, Rajpurkar P, Irvin J, Dunnmon J. PENet—a scalable deep-learning model for automated diagnosis of pulmonary embolism using volumetric CT imaging. npj Digital Medicine. 2020 Apr 24;3(1):1–9.

36. Kiani A, Uyumazturk B, Rajpurkar P, Wang A, Gao R, Jones E, Yu Y, Langlotz CP, Ball RL, Montine TJ, Martin BA. Impact of a deep learning assistant on the histopathologic classification of liver cancer. NPJ digital medicine. 2020 Feb 26;3(1):1–8.

37. Choi DJ, Park JJ, Ali T, Lee S. Artificial intelligence for the diagnosis of heart failure. NPJ Digital Medicine. 2020 Apr 8;3(1):1–6.

38. Riley RD, Ensor J, Snell KI, Harrell FE, Martin GP, Reitsma JB, Moons KG, Collins G, van Smeden M. Calculating the sample size required for developing a clinical prediction model. Bmj. 2020 Mar 18;368.

