## supplemental appendices for "Non-invasive Diagnosis of Deep Vein Thrombosis from Ultrasound with Machine Learning"

**Supplemental Materials**

**Appendix A: details about algorithm training and internal validation data characteristics**

The ML model's task is to annotate vessels, find anatomical landmarks, and analyse vessel compression state automatically. DVT diagnosis is done by automatization of the standard clinical ultrasound compression algorithm in a heuristic computer programme, based on the biometrics acquired from the ML model during the scan. Thus, the ML model has been trained mainly on data from healthy volunteers (n=246, age range 18-84, BMI < 30) and compression sequences from consented patients with confirmed DVT (n=9). An overview over the inclusion criteria is given in Figure A1 and the training data population in Table A1. An overview over the internal validation set is shown in Table A2.

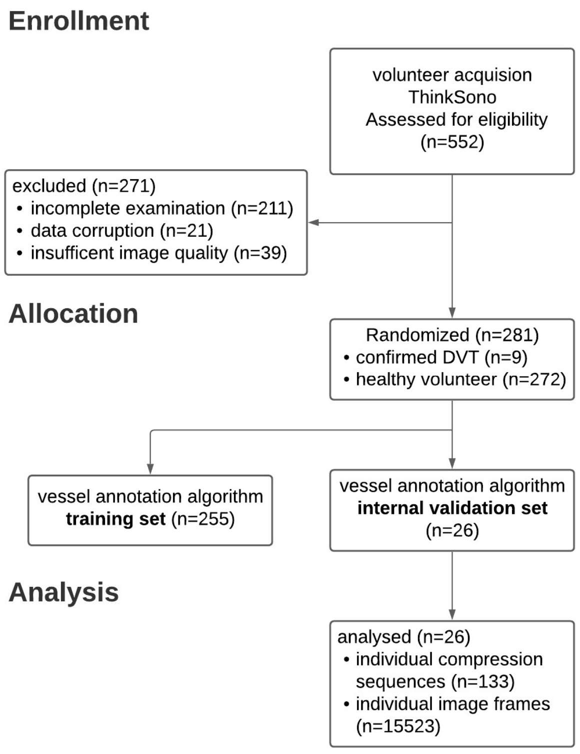

**Figure A1:** Consort diagram for inclusion of volunteer scans into the *training set* and *internal validation set*.

| **Algorithm Training data** | groin/thigh area model training | knee area model training | acquired data |
|---|---|---|---|
| subjects | 245 | 163 | 255 |
| number of compression sequences | 1076 | 616 | 1500 |
| **annotated scan sequences** | | | |
| background/no anatomical landmark or compression | 169 | 169 | 169 |
| LM0 - external iliac vein | 10 | - | 10 |
| *start of groin area after the inguinal ligament* | | | |
| LM1 - Greater saphenous vein + common femoral vein at saphenofemoral junction | 215 | - | 215 |
| LM2 - common femoral vein and artery | 51 | - | 51 |
| LM3 - common femoral vein and superficial and deep femoral arteries | 294 | - | 294 |
| LM4 - superficial and deep femoral veins and arteries | 141 | - | 141 |
| *end of groin area and beginning of thigh area at entrance to adductor canal* | | | |
| LM5 - proximal thigh with superficial vein clearly visible with deep femoral vein clearly separated in deep tissue | 123 | - | 123 |
| LM6 - mid thigh with superficial femoral vein and artery in the adductor canal | 288 | - | 288 |
| LM7 - distal thigh, same anatomy as LM6 | 2 | - | 2 |
| *end of thigh area and beginning of knee area* | | | |
| LM8 - proximal popliteal area, with popliteal vein and artery | - | 130 | 130 |

| | | | |
|---|---|---|---|
| LM9 - middle popliteal area, with tibial-fibular trunk and popliteal artery | - | 141 | 141 |
| LM10 - distal popliteal area, with anterior and posterior tibial and fibular veins and popliteal artery | - | 186 | 186 |
| Total number of manually annotated frames | 111546 | 88823 | 167145 |

**Table A1:** training data overview. Subjects may contain more than one landmark; thus, subject IDs may be present in training and internal validation set. Individual sequences are either in one or the other set. Landmarks used for the groin model in this study are highlighted orange and those for the knee area are green.

| **Internal validation data** | groin/thigh area validation sequences | knee area validation sequences | acquired internal validation sequences |
|---|---|---|---|
| total subjects | 25 | 17 | 26 |
| number of compression sequences | 88 | 58 | 133 |
| **annotated scan sequences** | | | |
| background/no anatomical landmark or compression | 13 | 13 | 13 |
| LM0 - external iliac vein | - | - | - |
| *start of groin area after the inguinal ligament* | | | |
| LM1 - Greater saphenous vein + common femoral vein at saphenofemoral junction | 17 | - | 17 |
| LM2 - common femoral vein and artery | 1 | - | 1 |
| LM3 - common femoral vein and superficial and deep femoral arteries | 27 | - | 27 |
| LM4 - superficial and deep femoral veins and arteries | 11 | - | 11 |
| *end of groin area and beginning of thigh area at entrance to adductor canal* | | | |

| | | | |
|---|---|---|---|
| LM5 - proximal thigh with superficial vein clearly visible with deep femoral vein clearly separated in deep tissue | 10 | - | 10 |
| LM6 - mid thigh with superficial femoral vein and artery in the adductor canal | 25 | - | 25 |
| LM7 - distal thigh, same anatomy as LM6 | - | - | - |
| *end of thigh area and beginning of knee area* | | | |
| LM8 - proximal popliteal area, with popliteal vein and artery | - | 20 | 20 |
| LM9 - middle popliteal area, with tibial-fibular trunk and popliteal artery | - | 11 | 11 |
| LM10 - distal popliteal area, with anterior and posterior tibial and fibular veins and popliteal artery | - | 16 | 16 |
| Total number of manually annotated frames | 9598 | 8257 | 15523 |

**Table A2**: The internal validation set represents a random 10% split at subject level of the overall available training data. Landmarks used for the groin model in this study are highlighted orange and those for the knee area are green.

**Appendix B: Details about the cost effectiveness model**

Test characteristics have been taken from[12] and are presented in Table B1 with the statistical distributions used in stochastic analysis presented in Tables B2-5.

| Description | | Mean |
|---|---|---|
| Wells Score: Proportion of patients with a proximal DVT characterised as: | | |
| | High risk | 0.68 |
| | Moderate risk | 0.25 |
| | Low risk | 0.07 |
| Wells Score: Proportion of patients without a proximal DVT characterised as: | | |
| | High risk | 0.11 |
| | Moderate risk | 0.41 |
| | Low risk | 0.48 |
| D-dimer (assumed to be an enzyme-linked immunosorbent assay (ELISA) test) | | |
| | Sensitivity for proximal DVT | 0.98 |
| | Specificity for proximal DVT when | |
| | Wells Score is high risk | 0.34 |
| | Wells Score is moderate risk | 0.45 |
| | Wells Score is low risk | 0.52 |
| Proximal ultrasound | | |
| | Test sensitivity | 0.95 |
| | Test specificity | 0.94 |

**Table B1**: Efficiency parameters for each test

| Description | Mean | Distribution | Parameter 1 | Source |
|---|---|---|---|---|
| Proportion of patients with proximal DVT characterised as high risk | 0.68 | Dirichlet | 105.61 | (Goodacre S., 2005) |
| Proportion of patients with proximal DVT characterised as moderate risk | 0.25 | Dirichlet | 38.83 | (Goodacre S., 2005) |
| Proportion of patients with proximal DVT characterised as low risk | 0.07 | Dirichlet | 10.87 | (Goodacre S., 2005) |
| Proportion of patients without proximal DVT characterised as high risk | 0.11 | Dirichlet | 40.78 | (Goodacre S., 2005) |
| Proportion of patients without proximal DVT characterised as moderate risk | 0.41 | Dirichlet | 151.99 | (Goodacre S., 2005) |
| Proportion of patients without proximal DVT characterised as low risk | 0.48 | Dirichlet | 177.94 | (Goodacre S., 2005) |

**Table B2**: Test Efficiency Well's test

| Description | Mean | Distribution | Param. 1 | Param. 2 | Source |
|---|---|---|---|---|---|

| | | | | | |
|---|---|---|---|---|---|
| Sensitivity for proximal DVT | 0.98 | Beta | 736.91 | 15.04 | (Goodacre S., 2005) |
| Specificity for proximal DVT - Wells's outcome: high risk | 0.34 | Fixed | | | (Goodacre S., 2005) |
| Specificity for proximal DVT - Wells's outcome: moderate risk | 0.45 | Beta | 4278.13 | 5228.83 | (Goodacre S., 2005) |
| Specificity for proximal DVT - Wells's outcome: low risk | 0.52 | Fixed | | | (Goodacre S., 2005) |

**Table B3**: Test Efficiency D-dimer test

| **Description** | | **Mean** | **Distrib.** | **Param. 1** | **Param. 2** | **Source** |
|---|---|---|---|---|---|---|
| Proximal DVT prevalence | | 0.147 | Beta | 41.00 | 238 | [22] |
| Treated proximal DVT | | | | | | |
| | Probability of fatal pulmonary embolus | 0.004 | Beta | 17.00 | 4204.00 | [12] |
| | Probability of non-fatal pulmonary embolus | 0.008 | Beta | 33.40 | 4070.60 | [12] |
| | Probability of post thrombotic syndrome | 0.053 | Beta | 28.00 | 500.00 | [12] |
| Outcomes associated with warfarin treatment | | | | | | |
| | Fatal haemorrhage | 0.003 | Dirichlet | a = 37 | | [12] |
| | Non-fatal intracranial haemorrhage | 0.001 | Dirichlet | b = 13 | | [12] |
| | Non-fatal non-intracranial haemorrhage | 0.021 | Dirichlet | c = 226 | | [12] |
| | No haemorrhage | 0.975 | Dirichlet | d = 10.481 | | [12] |
| Untreated proximal DVT | | | | | | |
| | Probability of fatal pulmonary embolus | 0.019 | Beta | 5.00 | 263.00 | [12] |
| | Probability of non-fatal pulmonary embolus | 0.093 | Beta | 25.00 | 243.00 | [12] |
| | Probability of post thrombotic syndrome | 0.330 | Beta | 5.21 | 10.57 | [12] |

**Table B4:** Proximal DVT prevalence and outcomes associated with treated and untreated proximal DVT

| Description | | Mean | Distribution | Parameter 1 | Parameter 2 | Source |
|---|---|---|---|---|---|---|
| Normal age-specific discounted quality adjusted life expectancy (QALYs) | | 11.58 | Fixed | | | [12] |
| Lifetime utility multiplier associated with | | | | | | |
| | Non-fatal pulmonary embolus | 0.940 | Beta | 19.43 | 1.24 | [12] |
| | Non-fatal intracranial haemorrhage | 0.290 | Beta | 8.34 | 20.41 | [12] |
| | Post thrombotic syndrome | 0.977 | Beta | 232.64 | 5.48 | [12] |
| Lifetime QALY's accrued by | | | | | | |
| | Patients with a DVT who are treated | 11.47 | [A] | | | |
| | Patients with a DVT who are untreated | 11.21 | [A] | | | |
| | Patients without a DVT who are treated | 11.54 | [A] | | | |

**Table B5:** QALYs associated with outcomes and QALYs accrued by patient category. [A]: The variance on these parameters is based on the variance of the parameters that make up these values

| Description | Mean [US $] | Distribution | Parameter 1 | Parameter 2 | Source |
|---|---|---|---|---|---|
| Well's test | 12.60 | Gamma | 25.00 | 0.34 | [12] |
| D-dimer | 33.20 | Gamma | 25.00 | 0.78 | [28] |
| Proximal ultrasound | 100.72 | Gamma | 319.23 | 0.25 | [29] |

**Table B6:** Costs of the diagnostic tests

Treatment reduces the probability of a patient with a DVT experiencing a fatal or non-fatal pulmonary embolism (PE) or post-thrombotic syndrome (PTS). However, treatment is associated with risks of fatal haemorrhage, non-fatal intracranial haemorrhage, and non-fatal non-intracranial haemorrhage.

According to[12] patients who do not experience any of a PE, PTS or a haemorrhage accrue a mean of 11.58 discounted lifetime QALYs. Mean quality of life multipliers for PTS, non-fatal PE and non-fatal intracranial haemorrhage of 0.977, 0.94 and 0.29 respectively were also presented by[12] with statistical distributions used in stochastic analysis presented in Tables B2-B5. These data were used to estimate total QALYs for the four diagnostic accuracy outcomes.

The lifetime, discounted, quality adjusted life years accrued by patients in each classification differ based on their true DVT status and their results from each diagnostic algorithm. Untreated patients with a DVT remain at high-risk of PE and PTS but do not have the risks of haemorrhage associated with treatment. Treated patients with a DVT have reduced risks of PE and PTS but have the risk of haemorrhage associated with treatment. Treated patients without a DVT have the same risk of PE and PTS as the general population but are subject to the risks of haemorrhage associated with treatment. Untreated patients without a true DVT will accrue the same discounted lifetime QALYs as the general population. The QALYs accrued in each of the four diagnostic accuracy outcomes are shown in Table B7.

| **Description** | **QALYs accrued** | **Cost incurred [US $]** |
|---|---|---|
| True Positive (DVT – treated) | 11 464 | 1 786 |
| False Negative (DVT – untreated) | 11 207 | 2 277 |
| False Positive (No DVT – treated) | 11 530 | 1 431 |
| True Negative (No DVT – untreated) | 11 580 | 0 |

**Table B7:** Estimated QALYs accrued and costs incurred for each diagnostic accuracy outcome. £ to $ conversion 1.34 as in 12/2020.

The discounted lifetime costs associated with patient outcomes were taken from[12,30]. Where appropriate costs were uplifted to 2018/19 values using inflation indices presented in[30].

The lifetime costs associated with PTS and non-fatal intracranial haemorrhage were both composite costs including the cost of a first attendance at a vascular surgery outpatient clinic and the cost of subsequent vascular surgery outpatient clinics visits for PTS and the cost of care in the first year and subsequent years in the case of non-fatal intracranial haemorrhage. The total cost associated with PTS and the method used to calculate these was included in[12] together with the costs of the components of the total cost. From this it was estimated that the expected lifetime of patients with PTS was 11.67 years. No such information was provided for patients experiencing a non-fatal, non-intracranial haemorrhage and thus it was assumed that the same expected lifetime applied when calculating costs.

Treatment for DVT consists of approximately eight days of low molecular weight (LMW) heparin followed by ninety days of warfarin. The total cost of DVT treatment of $1 102 is estimated using the same derivation as that used in[12] with one change: The current version of the British National Formulary[31] indicates that the initial dose of LMW heparin in the treatment of DVT is a large loading dose with subsequent smaller maintenance doses, thus the initial loading dose will be associated with a greater cost than subsequent maintenance doses. The costs of LWM heparin and warfarin are taken from the current version of the British National Formulary[31]. Additional resource use such as GP visits and anticoagulant clinic visits and their unit costs[12,30] and NHS Reference Costs 2015-2016[29], where appropriate cost have been inflated to 2018/19 values using inflation indices presented in[30]. The costs associated with outcomes associated with DVT or with treatment for DVT are shown in Table B8.

| **Description** | **Mean [US $]** |
|---|---|
| Treatment of fatal PE | 1 959 |
| Treatment of non-fatal PE | 1 901 |
| Lifetime treatment of PTS | 6 248 |
| Fatal intracranial haemorrhage | 11 082 |
| Lifetime treatment for non-fatal intracranial haemorrhage | 85 957 |
| Non-fatal non-intracranial haemorrhage | 955 |

**Table B8:** Costs associated with outcomes of DVT and complications associated with the treatment of DVT. £ to $ conversion 1.34 as in 12/2020.

The costs associated with each diagnostic test have been taken from[12,29] and are presented in Table B6.

**Appendix C: quality control scoring system**

We use a 10-point expert image quality scoring as it is outlined below to curate video data which has not been acquired under AutoDVT guidance and real-time quality control. The quality cut-off, i.e., the minimum required quality has been less or equal to a total score of 20 in this study.

1. Vessel boundaries in frame
    a. Fully in Frame: 1
    b. Cut off <50% of vessel size during parts sequence: 2
    c. Cut off <50% of vessel size during entire sequence: 3
    d. Cut off >50% of vessel size during parts of sequence: 4
    e. Cut off >75% of vessel size during part of sequence or >50% over entire sequence: 5
2. Adherence to regular LM configuration:
    a. Strongly adherent to LM configuration: 1
    b. Different positions of veins and arteries than regular configuration, mix of LMs: 2
    c. Loss of LM position for parts of sequence: 3
    d. Additional/missing (large) vessels for LM: 4
    e. Additional/missing vessels and different positions: 5
3. Contrast of veins to tissue
    a. Dark veins, bright tissue: 1
    b. Somewhat contrasted veins: 2
    c. Veins and tissue almost same echogenity: 3
4. Contrast of arteries to tissue:
    a. Dark arteries, bright tissue: 1
    b. Somewhat contrasted arteries: 2
    c. Arteries and tissue almost same echogenity: 3
5. Sharpness of vein boundaries
    a. Clear boundaries with strong dorsal echo amplification: 1
    b. Well discernible boundaries with some dorsal echo amplification: 2
    c. Poorly visible boundaries without echo amplification: 3
6. Sharpness of artery boundaries
    a. Clear boundaries with strong dorsal echo amplification: 1
    b. Well discernible boundaries with some dorsal echo amplification: 2
    c. Poorly visible boundaries without echo amplification: 3
7. Overall gain of image
    a. Medium gain range, good quality: 1
    b. Image too bright: 2
    c. Image too dark: 3
8. Depth of image
    a. Image ends about 1 cm below lowest vessel: 1
    b. Image ends within 1-2 cm below lowest vessel: 2
    c. Image ends 2+ cm below lowest vessel: 3
9. Image artefacts
    a. Good quality, only minor artefacts: 1
    b. Multiple smaller artefacts, also in/over the vessels: 2
    c. Large image problems, i.e., probe not fully on leg: 3
10. Probe Movement in sequence
    a. Medium paced compression and decompression, no lateral or horizontal movements, full vein compression: 1

b. Very fast or very slow compression and decompression, no lateral or horizontal movements, incomplete compression on healthy veins: 2
c. Minimal lateral/horizontal movement: 3
d. Lots of movement: 4

Total: 10-35 Points
